# Double-Quencher Probes Improved the Detection Sensitivity of Severe Acute Respiratory Syndrome Coronavirus 2 (SARS-CoV-2) by One-Step RT-PCR

**DOI:** 10.1101/2020.03.17.20037903

**Authors:** Yosuke Hirotsu, Hitoshi Mochizuki, Masao Omata

## Abstract

**Background:** Severe acute respiratory syndrome coronavirus 2 (SARS-CoV-2) emerges in Wuhan City, Hubei Province, spreads worldwide, and threats the human life. The detection of SARS-CoV-2 is important for the prevention of the outbreak and management of patients. Real-time reverse-transcription polymerase chain reaction (RT-PCR) assay detected the virus in clinical laboratory.

**Methods:** This study utilized primers and single-quencher probes in accordance with the Centers for Disease Control and Prevention (CDC) in the USA and the National Institute of Infectious Diseases (NIID) in Japan. Moreover, we designed the double-quencher probes (YCH assay) according to the oligonucleotide sequence established by NIID. Using these assays, we conducted a one-step real-time RT-PCR with serial DNA positive control to assess the detection sensitivity.

**Results:** The threshold cycle (Ct) value of RT-PCR was relatively low in CDC and YCH assays compared to NIID assay. Serial dilution assay showed that both CDC and YCH assays could detect a low-copy number of DNA positive control. The background fluorescent signal at the baseline was lower in YCH than that of NIID.

**Conclusion:** Double-quencher probes decreased background fluorescent signal and improved detection sensitivity of SARS-CoV-2.

## Introduction

In early December 2019, the first pneumonia cases of unknown origin were identified in Wuhan, the capital city of Hubei province [1]. The novel human-infecting coronavirus was tentatively named 2019 novel coronavirus (2019-nCoV) by the World Health Organization (WHO) and, currently, renamed as severe acute respiratory syndrome coronavirus 2 (SARS-CoV-2) by the International Committee on Taxonomy of Viruses. As of March 9, 2020, more than 109,577 cases of coronavirus disease 2019 (COVID-19) had been confirmed, of which 80,904 cases are in China and 28,673 cases outside of China, with 3,809 deaths globally (WHO Situation Report–49). Human-to-human transmission of the virus accounts for the spread of the virus around the world [2-7]. Human angiotensin-converting enzyme 2 (ACE2) is the receptor for SARS-CoV-2 and one of the potential therapeutic targets [8, 9].

SARS-CoV-2 belongs to a group of SARS-like coronaviruses (genus *Betacoronavirus*, subgenus *Sarbecovirus*) which had previously been found in bats in China [10]. Phylogenetic analysis indicated that bats are possibly the original hosts of this virus [11]. SARS-CoV-2 has a single-strand, positive-sense RNA genome with 29,903 base pairs in length (NCBI Reference Sequence: NC_045512.2) [12]. A viral genome sequence was released for immediate public health support via the community online resource (virological.org), followed by 169 genome-sequencing data that were deposited in the database curated by the Global Initiative on Sharing All Influenza Data (GISAID) [13]. These data showed how SARS-CoV-2 outbreaks occurred worldwide in genetic variations.

Human transmission of SARS-CoV-2 has become a global health concern. The molecular diagnosis of SARS-CoV-2 is highly prioritized to establish the prevention of the viral outbreak and treatment of infected patients. Therefore, demands in detecting the virus in suspected patients in hospitals and clinical laboratories are growing. The Japanese government announced the approval of real-time PCR analysis to detect SARS-CoV-2 for health insurance on March 6, 2020. Here, we compared the detection sensitivity among the primer/probe sets designed by the National Institute of Infectious Diseases (NIID) in Japan and the Centers for Disease Control and Prevention (CDC) in the United States of America. These probes were designed as single-quencher probes. We originally designed the double-quencher probes consisting of the same nucleotide sequencing imposed by the NIID. The results showed that double-quenching probes reduced the background signal and improved the detection sensitivity of SARS-CoV-2.

## Materials and Methods

### Primer and probe sets

The CDC has designed real-time reverse-transcription polymerase chain reaction (RT-PCR) assays and published a protocol for the detection of the 2019-nCoV (https://www.cdc.gov/coronavirus/2019-ncov/lab/rt-pcr-panel-primer-probes.html). We purchased the 2019-nCoV CDC qPCR Probe Assays (renamed as 2019-nCoV RUO Kit) from Integrated DNA Technologies (IDT, Coralville, IA, USA), which contained research-use-only primer and probe sets based on the protocol announced from CDC (hereafter called CDC assay). The CDC assay includes three sets of primers and 5’ FAM dye and 3’ Black Hole Quencher® (BHQ) probes (Table 1).

**Table 1.**
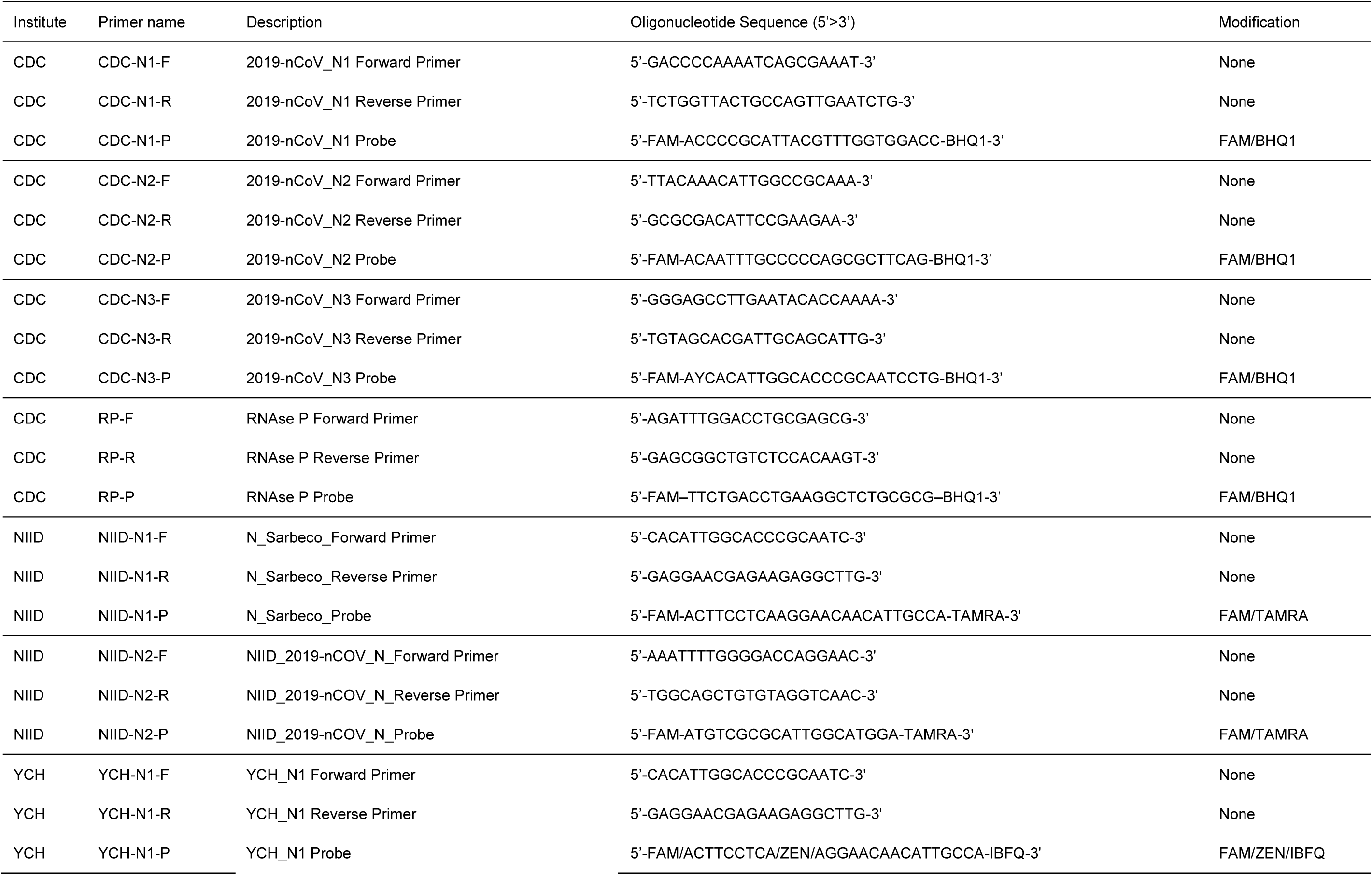
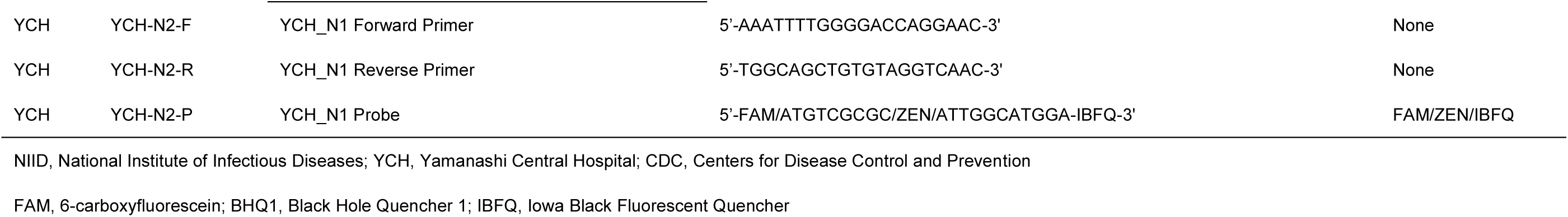
Sequences of the primers and probes used in the CDC, NIID, and YCH assays

The NIID has designed RT-PCR assays and published the data [14]. The primer and probe sets were sent from the NIID (hereafter called NIID assay). The CDC assay includes three sets of primers and 5’ FAM dye and 3’ TAMRA probes as shown in Table 1.

We also originally designed double-quencher probes (IDT) containing the same sequence in accordance with the NIID protocol (hereafter called YCH assay). This probe has 5’ FAM dye, internal ZEN Quencher, and 3’ Iowa Black® Fluorescent Quencher (IBFQ) (Table 1). Internal ZEN Quencher is incorporated between the ninth and tenth bases from the 5’ end of probe. This design decreased the distance between the dye and quencher and expected to reduce the background signal and achieve a higher dynamic signal. All primer/probe sets of CDC, NIID, and YCH assays amplified the nucleocapsid (N) gene of SARS-CoV-2 (Figure 1). CDC assay targets three sites of N gene, while both NIID and YCH target the two sites (Figure 1).

**Figure 1.**
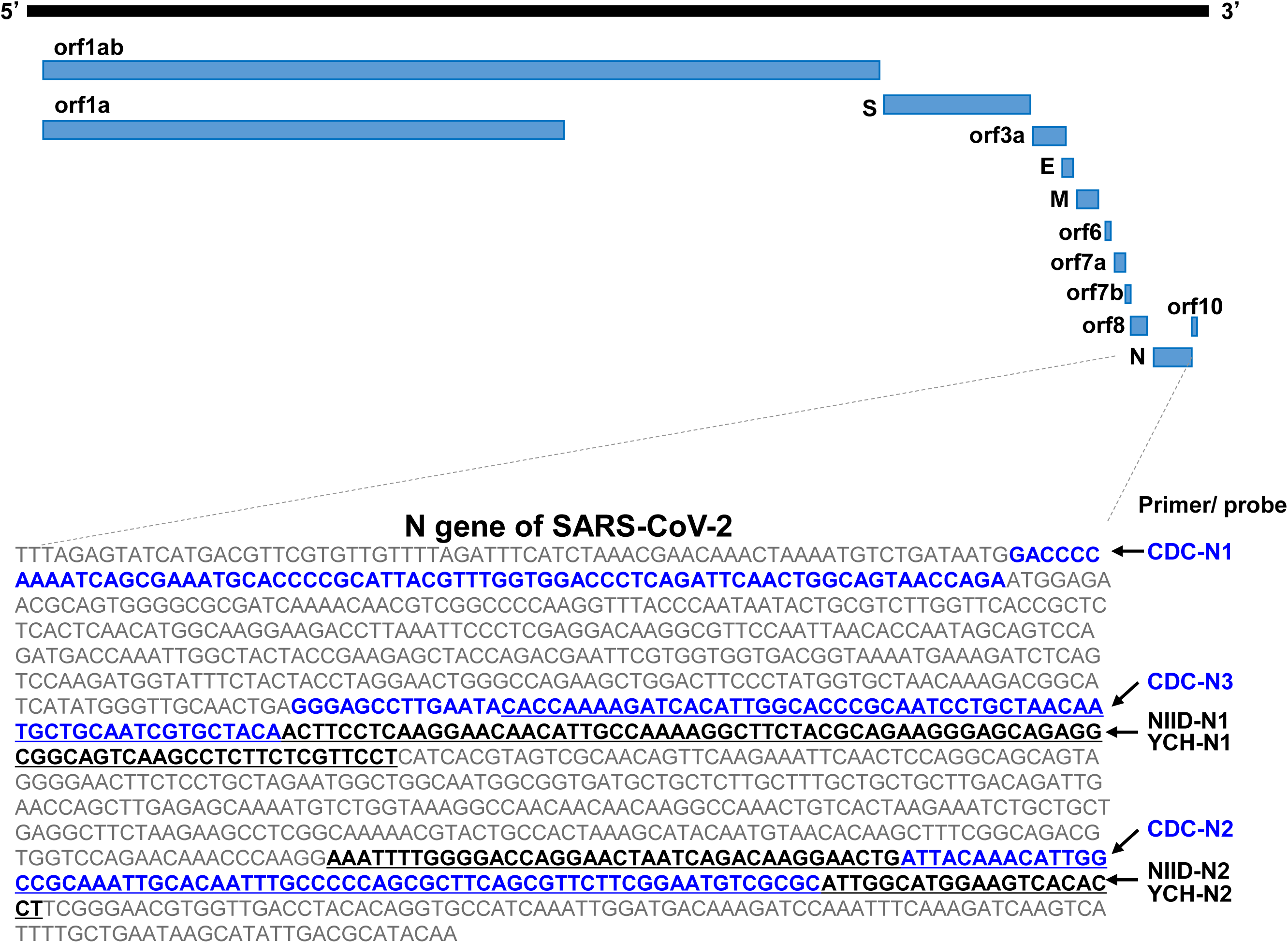
Genomic structure of SARS-CoV-2 and design of primer/probe. SARS-CoV-2 is a single-strand RNA virus, harboring 29,903 base pairs. Primer and probes were designed to nucleocapsid (N) protein gene. The CDC assay (USA) targets three sites in the N gene (CDC-N1, CDC-N2, CDC-N3). The NIID (Japan) and YCH target the same two sites (NIID-N1, YCH-N1, NIID-N2, and YCH-N2). The CDC and NIID assays are composed of single-quencher probe, whereas the YCH is composed of double-quencher probe.

### DNA positive control

For the DNA positive control, we purchased 2019-nCoV_N_Positive Control (IDT, catalog #10006625), which is composed of plasmids containing the complete N gene (1,260 base pairs) of SARS-CoV-2. We prepared a serial dilution DNA control using nuclease-free water (Thermo Fisher Scientific Waltham, MA, USA) to evaluate the detection sensitivity of CDC, NIID, and YCH assays. We also purchased Hs_RPP30 Positive Control (IDT, catalog #10006626), which contains a portion of the ribonuclease P 30 subunit (RPP30) gene of the human genome. RPP30 genes were analyzed for internal positive control.

### One-step real-time RT-PCR

To detect SARS-CoV-2, we performed RT-PCR with the use of probes and primers. In CDC and YCH assays, the reaction mixture was made up of 5 μL of 4× TaqMan Fast Virus 1-Step Master Mix (Thermo Fisher Scientific), 1.5 μL of probe/primer mixtures (IDT), 11.5 μL of nuclease-free water (Thermo Fisher Scientific), and 2 μL of serial-diluted DNA control (2019-nCoV_N_Positive Control) in a 20 μL total volume. We also performed one-step real-time PCR using the NIID assays according to the protocol (version 2.7) established by the NIID. Briefly, in the N1 of the NIID assay, the reaction mixture comprised of 5 μL of 4× TaqMan Fast Virus 1-Step Master Mix, 1.2 μL of forward primer, 1.6 μL of reverse primer, 0.8 μL of probe, 9.4 μL of nuclease-free water (Thermo Fisher Scientific), and 2 μL of serial-diluted DNA control in a 20 μL total volume. In the N2 of the NIID assay, the reaction mixture consisted of 5 μL of 4× TaqMan Fast Virus 1-Step Master Mix, 1.0 μL of forward primer, 1.4 μL of reverse primer, 0.8 μL of probe, 9.8 μL of nuclease-free water (Thermo Fisher Scientific), and 2 μL of serial-diluted DNA control in a 20 μL total volume. RT-PCR was conducted under a ViiA7 Real-Time PCR System (Thermo Fisher Scientific) with the following cycling conditions: 50°C for 5 min for reverse transcription, 95°C for 20 s, and 45 cycles of 95°C for 3 s and 60°C for 30 s. Data were analyzed using the ViiA7 software v2.2.2 (Thermo Fisher Scientific). The threshold line was set at 0.2. The threshold cycle (Ct) value assigned to each PCR reaction and amplification curve was visually assessed.

## Results

### Design of primer and probes to detect SARS-CoV-2

SARS-CoV-2 has a single-strand, positive-sense RNA genome with 29,903 base pairs in length (NCBI Reference Sequence: NC_045512.2). All CDC, NIID, and YCH assays amplified the N gene of the SARS-CoV-2, as shown in Figure 1. CDC assays target three sites (N1, N2, and N3), and NIID and YCH assays target two sites (N1 and N2) (Figure 1). Previous study showed the mismatch of reverse primer in the N1 of the NIID assay with the sequence in the current database because it was constructed based on another reported sequence (GenBank: MN908947.1) [14]. In addition, this mismatch in the reverse primers had no influence on the detection sensitivity compared to the perfect-matched reverse primer stated in the previous report [14]. CDC and NIID assays contain single-quencher probes in each assay; however, we originally generated the double-quencher technology in YCH assays to reduce background signal. Expected amplicon sizes of CDC assays are 72 bp, 67 bp, and 72 bp in length by the N1, N2, and N3, respectively. Expected amplicon sizes of NIID and YCH assays are 128 bp and 158 bp in length by N1 and N2, respectively.

### Serial dilution analysis determined detection sensitivity

To assess the detection sensitivity of each assay, we conducted RT-PCR analysis using a serial-diluted DNA control. We input the DNA positive control at concentrations of 10,000 copies, 1,000 copies, 100 copies, 10 copies, and 1 copy. We set the threshold value to 0.2 in all assays to determine the threshold cycle (Ct). Fluorescence amplification signals were detected using 10,000 copies, 1000 copies, and 100 copies of DNA positive control by all CDC, NIID, and YCH assays (Figure 2). With the use of 10 copies of DNA positive control, amplification signals by CDC-N2, YCH-N1, YCH-N2, and NIID-N2 were detected (Figure 2). All of these assays could not detect using 1 copy of DNA positive control (Figure 2). Overall, the Ct value is found to be lower in the YCH assay using the serial-diluted DNA positive control.

**Figure 2.**
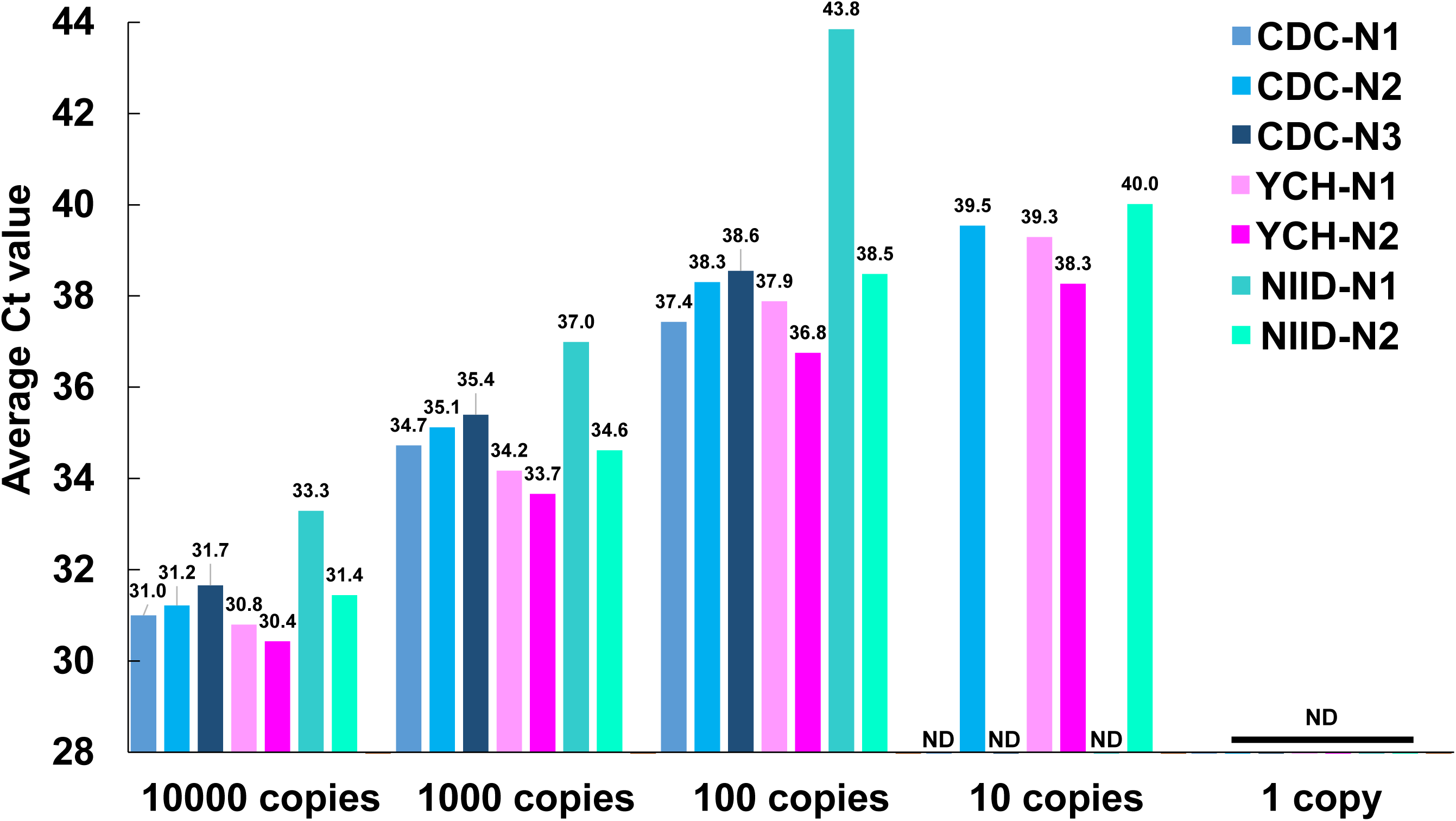
Serial dilution assays determined the detection sensitivity of SARS-CoV-2. The DNA positive control was diluted from 10,000 copies to 1 copy. The threshold value was set as 0.2. Bar plot shows the average threshold cycle (Ct) in each assay. The number above the bar is the average Ct value. ND, not detected.

### Double-quencher probes reduce background signal

As observed, the Ct values in both the N1 and N2 sites of the NIID assay were relatively higher compared to those of the YCH assay (Figure 2), although the primer and probe sequence was the same between NIID and YCH assays. We concluded that the double-quencher probe (ZEN/IBFQ) reduced the background signal and lowered the Ct value compared to the single-quencher probe (TAMRA). To this end, we analyzed the fluorescent signals during the cycles of RT-PCR at three independent experiments. As expected, the double-quencher probes decreased the background FAM fluorescent signals at the baseline (Figure 3). The baseline fluorescent signal of the N1 site of NIID ranged from 150×10^4^ to 200×10^4^. However that of YCH was around or less than 50×10^4^ (Figure 3). Similarly, the signal in the N2 site of NIID was around 125×10^4^ to 200×10^4^, and that of YCH was 75×10^4^ to 100×10^4^. Results suggested that double-quencher probes improved the detection sensitivity of SARS-CoV-2 using one-step real-time RT-PCR owing to the reduction of background signal.

**Figure 3.**
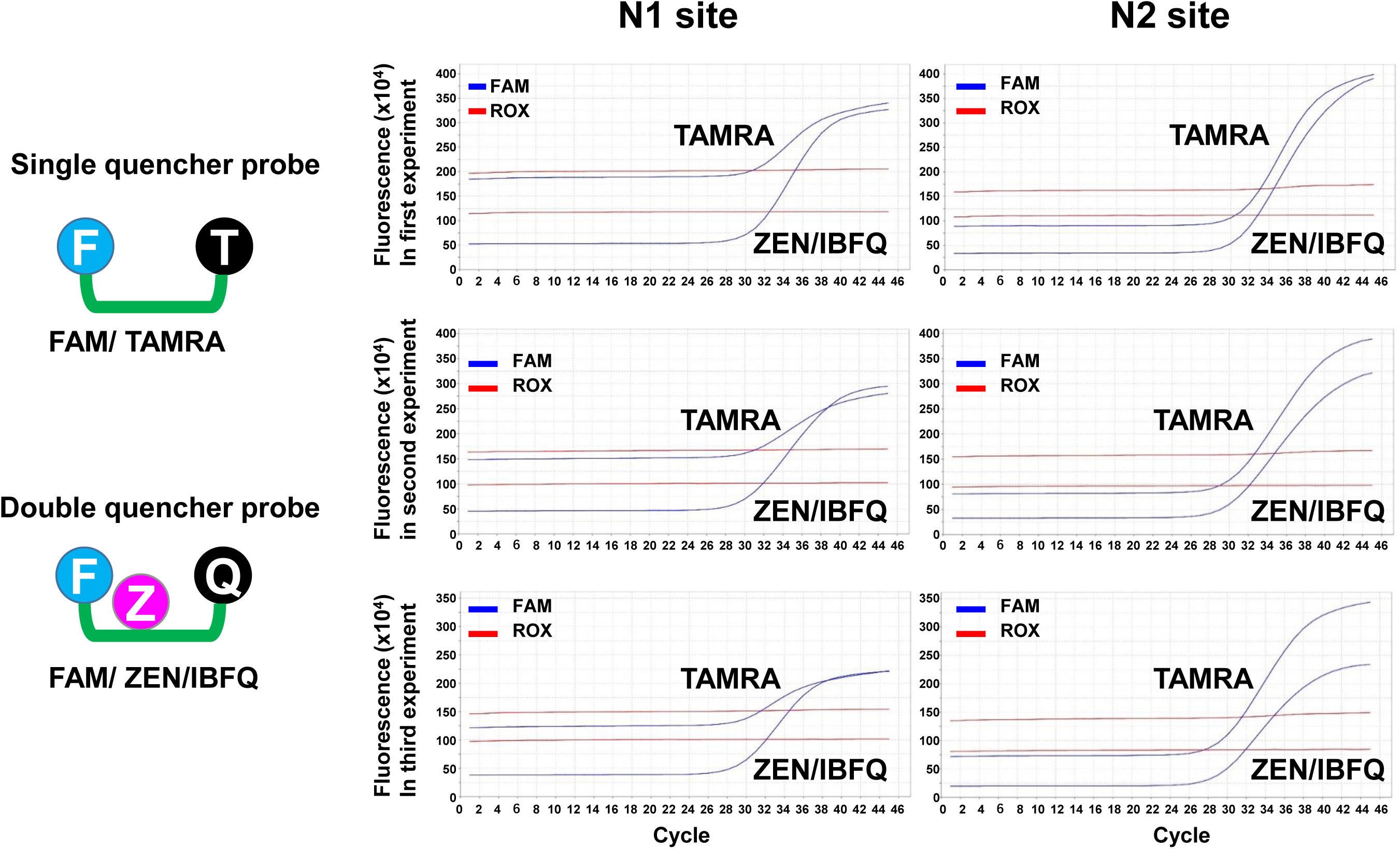
Double-quencher reduce the background fluorescent signal. The right image shows the single- or double-quencher probe. Single-quencher probe has FAM dye and TAMRA quencher (NIID assay). Double-quencher probe has FAM dye, ZEN internal quencher, and IBFQ quencher (YCH assay). Experiment was independently conducted three times. Blue line shows FAM signal and red shows ROX signal. ZEN/IBFQ double-quencher reduced the background fluorescent signal compared to the TAMRA single-quencher in both N1 and N2 sites.

## Discussion

SARS-CoV-2 is the primary cause of an ongoing international outbreak of a respiratory illness known as COVID-19. The detection of this virus is important in order to prevent outbreak and manage patient care. In this study, we performed RT-PCR using different types of primer and probes. As a result, double-quencher probes decreased background signals compared to single-quencher probes. Owning to less background signals, the dynamic changes of a single fluorescence is high and demonstrate a lower Ct value. Results showed the relevance of a double-quencher probe in increasing the sensitivity in the detection of SARS-CoV-2.

Several companies developed a commercially available kit for the RT-PCR. To the best of our knowledge, the IDT launched assays targeting the three sites of N gene and human RPP30, as used in this study. Thermo Fisher Scientific provided the assay targeting Orf1ab, spike (S) gene, N gene, and human RNase P; Roche Diagnostics provided an assay targeting RdRP (in Orf1ab), E gene, and N gene. Jung *et al*. recently compared several types of primer and probe sets reported in China, Germany, Hong Kong, Japan, Thailand, and the USA [15]. They showed that ORF1ab is best according to China. In the N gene, CDC-N2 (USA), CDC-N3 (USA), and NIID-N2 (Japan) sets should be selected for sensitive and reliable laboratory confirmation of SARS-CoV-2. We have also observed that the CDC-N2 (USA) and NIID-N2 (Japan) assay could detect low number of DNA positive control.

We confirmed that the N1 site of NIID assay was not detected when using less than 10 copies of DNA positive control. In addition, as observed, Ct value was also high by the N1 site of the NIID assay. Similar to these observations, Shirato *et al*. also demonstrated less sensitivity of the N1 site of NIID assay using RNA positive control. The assessment of N1 site was originally reported by the groups in Germany. To improve the detection sensitivity, we recommended the use of a double-quencher probe for detection of SARS-CoV-2, especially in the N1 site.

The background fluorescent signal was mainly determined by two factors. First, high background fluorescence is due to longer probe sequence in length. This means the shorter the distance between the fluorescence dye and the quencher, the better the quenching performance due to the fluorescence resonance energy transfer. Second, the background signal is prone to be low if the probe forms three-dimensional structure owing to self-quenching. To detect the low number of virus in human sample, the reduction of background signal is important. To this end, we utilized a double-quencher system for detecting SARS-CoV-2 by a one-step real-time RT-PCR. As expected, a double-quencher probe decreased the background signal at the baseline which led to a higher dynamic range. Our data suggested that double-quencher probe enables us to detect low number of SARS-CoV-2 in routine clinical practice.

## Data Availability

Data is available upon request.

## Acknowledgments

We thank all of the medical and ancillary hospital staff and the patients for consenting to participate. This study was supported by a Grant-in-Aid for the Genome Research Project from Yamanashi Prefecture (to M.O. and Y.H.), the Japan Society for the Promotion of Science (JSPS) KAKENHI Early-Career Scientists (grant number JP18K16292 to Y.H.), a Research Grant for Young Scholars (to Y.H.), the YASUDA Medical Foundation (to Y.H.), the Uehara Memorial Foundation (to Y.H.), and Medical Research Grants from the Takeda Science Foundation (to Y.H.).

